# Near real-time surveillance of the SARS-CoV-2 epidemic with incomplete data

**DOI:** 10.1101/2021.01.25.20230094

**Authors:** PM De Salazar, F Lu, JA Hay, D Gómez-Barroso, P Fernández-Navarro, E Martínez, J Astray-Mochales, R Amillategui, A García-Fulgueiras, MD Chirlaque, A Sánchez-Migallón, A Larrauri, MJ Sierra, M Lipsitch, F Simón, M Santillana, MA Hernán

## Abstract

Designing public health responses to outbreaks requires close monitoring of population-level health indicators in real-time. Thus, an accurate estimation of the epidemic curve is critical. We propose an approach to reconstruct epidemic curves in near real time. We apply this approach to characterize the early SARS-CoV-2 outbreak in two Spanish regions between March and April 2020.

We address two data collection problems that affected the reliability of the available real-time epidemiological data, namely, the frequent missing information documenting when a patient first experienced symptoms, and the frequent retrospective revision of historical information (including right censoring). This is done by using a novel back-calculating procedure based on imputing patients’ dates of symptom onset from reported cases, according to a dynamically-estimated “backward” reporting delay conditional distribution, and adjusting for right censoring using an existing package, *NobBS*, to estimate in real time (nowcast) cases by date of symptom onset. This process allows us to obtain an approximation of the time-varying reproduction number (*R*_*t*_) in real-time.

At each step, we evaluate how different assumptions affect the recovered epidemiological events and compare the proposed approach to the alternative procedure of merely using curves of case counts, by report day, to characterize the time-evolution of the outbreak. Finally, we assess how these real-time estimates compare with subsequently documented epidemiological information that is considered more reliable and complete that became available later in time. Our approach may help improve accuracy, quantify uncertainty, and evaluate frequently unstated assumptions when recovering the epidemic curves from limited data obtained from public health surveillance systems in other locations.

## Introduction

Assessing the effectiveness of ongoing interventions during outbreaks requires a real-time characterization of new infections. Epidemic curves, a monitoring tool frequently proposed to characterize the dynamics of an outbreak, capture the number of individuals infected over time. Reconstructing these curves appropriately is challenging for novel pathogens because testing protocols and surveillance systems may not be repurposed quickly enough.

Ideally, epidemic curves should document new infections based on the date of exposure to the infectious agent for each individual [1]. Because information on the date of exposure for each patient is usually unavailable, epidemic curves are often reconstructed based on the first detectable clinical event: onset of symptoms.

In practice, however, detected cases are rarely documented at onset of symptoms. Rather, surveillance procedures tend to rely on confirmed diagnoses, which are typically reported with a delay of days or weeks after symptoms onset [2–4]. As a result, the number of cases based on the onset of symptoms in any given day is unknown until those cases are reported days or weeks later. This problem is sometimes referred to as “backfill bias”, a term from economics [5] that was later applied to infectious disease tracking [6,7].

Reporting delays complicate timely decision-making [8]. Further, similar populations with different notification procedures may have different reporting delays [9] and the distribution of delays may change over time [10,11]. One way to address the problem of delayed notification is to statistically predict the number of cases with onset of symptoms today based on historical observations of how the number of cases on a given day were later revised to reflect updated information. This methodology is referred to as “nowcasting” [6,10]. A recently proposed approach to nowcasting, *NobBS* or Nowcasting by Bayesian Smoothing [10], complements the estimated delay distribution and historical data with the intrinsic autocorrelation from the transmission process. *NobBS* has been shown to perform better than previous nowcasting methods for different infectious diseases [10].

Nowcasting requires that the date of symptom onset is collected for all cases that are eventually reported. However, real-time surveillance systems cannot guarantee the ascertainment of the date of the clinical event for all reported cases, even if all cases are reported. Therefore, nowcasting the epidemic curve based on symptoms onset requires imputation of the missing dates of symptoms onset.

The nowcast epidemic curve can then be used to estimate the time-varying reproductive number (*R*_*t*_), that is, the number of secondary infections arising from a single infection on a particular day [12,13]. The estimation of the reproductive number requires also parametric assumptions on the generation interval (the time between a primary and a secondary infection).

Here, we present a three-step approach using existing computational tools to estimate in near-real time the epidemic curve and the time-varying reproductive number (*R*_*t*_) in the presence of reporting delays and incomplete data on the date of onset of symptoms. First, we impute the missing date of onset of symptoms data using historical distributions derived from reported line-list data. Second, we use *NobBS* to estimate case counts up to the present while adjusting for reporting delays. Third, we estimate the time-varying reproductive number using the nowcasted epidemic curve. We apply the approach to data reported during the early stages of the SARS-CoV-2 outbreak in two regions of Spain.

## Methods

### Surveillance data

We applied our methodology to Madrid and Murcia, two regions of Spain with very different characteristics. Madrid has 6.7 million residents, is highly interconnected both nationally and internationally, has the highest population density and urbanicity of the country, is situated in the geographic (inland) center, and had a seroprevalence for SARS-CoV-2 of 11.5% at the end of the study period [14]. Murcia has 1.4 million residents, average connectivity, population density and urbanicity, is geographically situated in the coastal periphery and had a seroprevalence of 1.6% at the end of the study period.

Each region reported daily counts of PCR-confirmed COVID-19 cases to the Spanish Ministry of Health [15]. We also obtained individualized data on date of report (DOR) and, for a proportion of cases, the date of symptoms onset (DOS) from the Spanish System for Surveillance at the National Center of Epidemiology (RENAVE) through the Web platform SiViEs (System for Surveillance in Spain) [16].

We conducted the analyses in each region using cumulative data available at three overlapping periods in the outbreak: early analysis period when reported cases reached maximum counts (spanning March 1-March 27), intermediate analysis period shortly after the peak of the epidemic curve (March 1-April 9), and late analysis period when the epidemic curve was close to zero (March 1-April 16). All analyses were implemented in R.

### Step 1: Imputation of missing data

We imputed the missing DOS by randomly assigning values drawn from the distribution of reporting delay (the period between DOS and DOR) in individuals with known DOS. Of note, the reporting delay distribution conditional on the DOR is different from both the unconditional delay distribution (the distribution of all reporting delays) and the “forward” delay distribution conditional on the DOS (*if my DOS is today, how long do I wait until DOR?*). Inferring reporting delays from the unconditional delay distribution or the “forward” delay distribution conditional on the DOS is known to generate biased reconstructed epidemic curves [17].

We assumed that missing DOS occur at random within region *i* and report date *t* and that reporting delays conditional on DOR can be modeled as a negative binomial with mean parameter *μ*_*i,t*_ and dispersion parameter *θ*_*i*_ using maximum likelihood [9]. We resampled 100 times to generate 100 time series of cases with complete DOS-DOR for each region *i*. The sum of the observed and imputed cases became the total cases used for nowcasting.

In sensitivity analyses, we first evaluated the main procedure after: a) masking DOS (hiding original data as missing values) in a random 10% of reported cases, b) masking DOS in a random 40% of reported cases, and c) masking DOS in a random 40% of cases as in b), then simply subtracting the observed mean delay to the reported date, resulting in the same imputed day for all cases reported at any given *t* [17].

### Step 2: Nowcasting the epidemic curve

After imputation, all reported cases have a DOS. However, a number of cases with DOS before day *t* will be reported after day *t*. We therefore used NobBS [10] to nowcast the number of yet unreported cases with DOS at *t* that would be eventually reported under the assumption that the delay conditional on DOS occurs as a negative binomial process. Briefly, *NobBS* uses historical information on the reporting delay to predict the number of not-yet-reported cases in the present using a log-linear model of the number of cases. The implementation of *NobBS* requires the specification of 1) a sliding window for the time-varying reporting delay, 2) a maximum delay allowed for the window, and 3) a prior distribution on the expected mean as a function of the “true” epidemiological signal (here, the number of new cases with onset of symptoms) *α*_*t*_ and the probability of delay *β*_*d*_. We used a moving window that represented 75% of the total period of analysis and a uniform prior distribution for the parameters *α*_*t*_ and *β*_*d*_. The NobBS method can be implemented in R using the *NobBs* package (v1.2), which compiles in *JAGS* using the *rjags* package (v4.10).

To assess uncertainty in the estimation, we produced a nowcast series with 10,000 posterior samples for each time point of the 100 imputed case count series in each region. We then pooled the samples and calculated the nowcast median and 2.5 and 97.5 percentiles for each day, following the recommendation of Zhou and Reiter [18].

In sensitivity analyses, we varied the length of the window using the period comprising 50% or 99% of the last date of report delays, and we generated nowcasts using the alternative model proposed in sensitivity Step 1c (using cases by report day and backshift by the mean reporting delay).

### Step 3: Estimation of time-varying reproductive number *R*_*t*_

We estimated the time-varying reproduction number *R*_*t*_ using two approaches implemented in the R package *epiEstim* (v2.2.1): Wallinga and Teunis [13,19]) (WT) and Cori et al. (C) [13,19]. We used the nowcast estimates and a mean generation interval (the time between a primary and a secondary case infection) of 5 days with a standard deviation of 1.9 [20]. We computed *R*_*t*_ from the 10,000 *NobBS* samples to produce the mean and 95% credibility interval of the *R*_*t*_ (for computational ease, we used a random sample of size 100 from the 10,000 samples since results did not materially change with a greater sample). Because WT and C use different forward and backward-looking computational approaches, respectively, a relative delay approximating the mean generation interval of WT relative to C is to be expected [17].

In sensitivity analyses, we used a significantly longer generation interval (mean=7.5, SD=3.5) [21], and also used an epidemic curve generated using observed cases by DOR backshifted by the mean reported delay.

## Results

### Imputation of missing data

The proportion of missing DOS among reported cases varied by region and epidemic phase. In Madrid, the percentage of missingness was 53% of 32,723 reported cases in the early analysis period, 37% of 50,745 in the intermediate analysis period, and 16% of 56,057 in the late analysis period. The corresponding numbers in Murcia were 25% of 831, 23% of 1433, and 11% of 1602. The distribution of the reporting delay was also region-specific and changed over time (Supplementary material, Figure S1).

The first row of Figure 1 for each region shows the weekly counts of cases by DOR with observed and missing DOS. The second row of Figure 1 shows the epidemic curve by DOS after the median of imputed cases (grey) was added to the observed cases with available DOS (blue). As expected, the uncertainty of the imputation increases with the proportion of missingness (Supplementary material, Figure S2).

**Figure 1.**
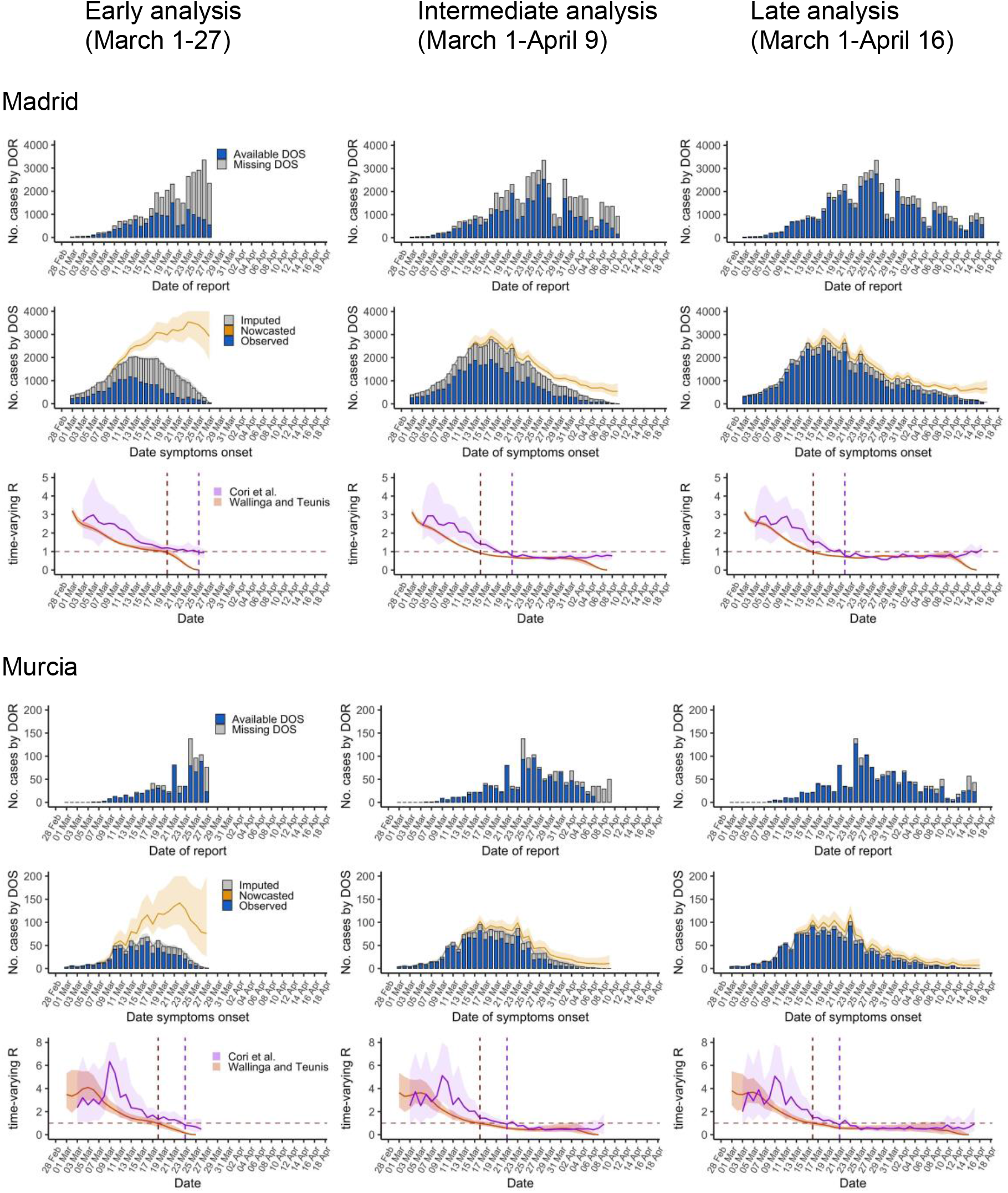
Epidemic curves and reproductive numbers estimated using the data available during the early, intermediate, and late analysis of the initial SARS-CoV-2 outbreak in the regions of Madrid and Murcia, Spain, March 1-April 16, 2020 DOS: date of onset of symptoms; DOR: date of report; Lines are median estimates, ribbons span 2.5 and 97.5 percentiles. Vertical lines indicate the day when *R*_*t*_ <1 (red dashed line for WT, purple dashed line for C)

### Nowcasting the epidemic curve

The second row of Figure 1 shows the epidemiologic curve after nowcasting (yellow line). Nowcasting reconstructed the epidemic curve more reliably than unadjusted case counts, either by DOR or DOS, even in earlier phases. This is more easily seen in Figure 2: nowcasted case counts in the intermediate period represent more accurately those estimated in the latest period, either compared with the curve of raw observed (not missing) case counts (B and E) or the curve modeled by mean backshift (C and F). However, in the early period of analysis for Madrid, with a high proportion of missing data, nowcasting overestimated those observed later in time.

**Figure 2.**
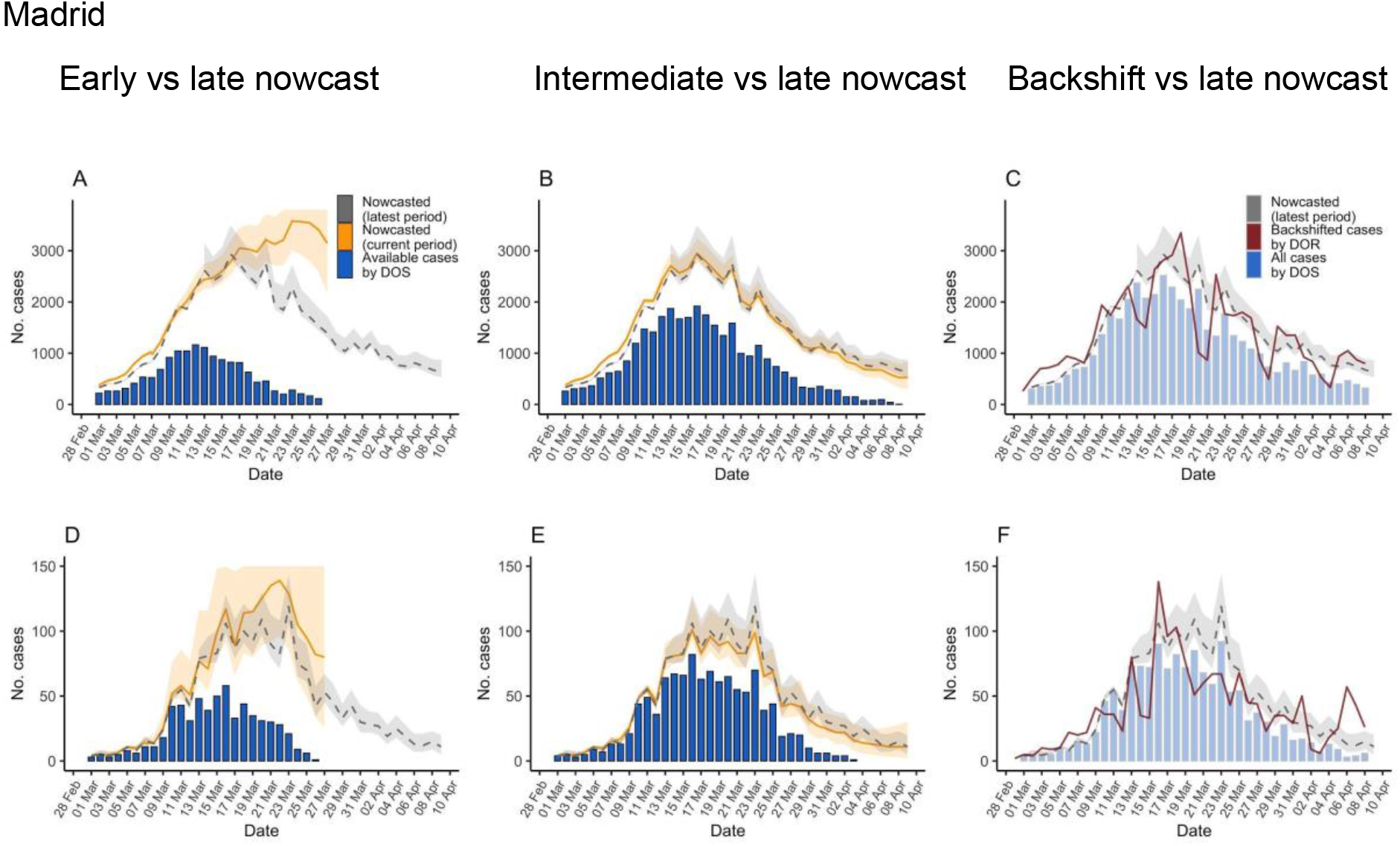
Epidemic curves estimated using the data available during the early and intermediate analysis of the initial SARS-CoV-2 outbreak in the regions of Madrid and Murcia, Spain, March 1-April 9, 2020, and comparison with curves obtained in late period of analysis March 1-April 16. Showing nowcast estimates (orange) in Madrid and Murcia for the early (A and D) and intermediate (B and E) period analysis, observed cases with known date of onset of symptoms for the same period (blue columns), and for comparison with more complete data, those estimated in the late period of analysis (dashed grey line and ribbon); C and F showing cases by date of report back shifted by the mean delay (red line) together with nowcast estimates for the late period analysis and observed cases with known date of onset of symptoms (shadowed blue columns). DOS: date of onset of symptoms; DOR: date of report; Lines are median estimates, ribbons span 2.5 and 97.5 percentiles.

Nowcast curves are smoother than curves of case counts by report date because of removal of noise related to the reporting process (such as weekday dependency). Also, the peak of case counts by symptom onset was several days earlier in the nowcast curves (around March 16 in both regions) than in the curves of reported cases (around March 25-26). The nowcasted peak is more consistent with the implementation of the country-wide lockdown by March 15 [22] (see Discussion). The uncertainty in the nowcast increases with uncertainty in the imputation of DOS, as can be seen in the large uncertainty band in the early analysis in Madrid.

The nowcast curves were relatively robust to missingness and different parameterizations of the *NobBS* function, such as window of analysis. However, the time of peak of case-counts can significantly vary with data availability and assumptions (Supplementary material, Figure S2).

### Estimation of the time-varying reproductive number

The third row of Figure 1 shows the *R*_*t*_ estimates using the nowcasted curve and including uncertainty from previous steps. The precision of *R*_*t*_ increased over time when more information became available, as seen in the third row of Figure 1 for both regions. Estimates using nowcasted curves show an earlier reduction of *R*_*t*_ than those obtained from raw case counts by DOR (Supplementary material, Figure S5) which mirrors the difference between the nowcasted curve, and the observed curve of case counts by report date. Though the WT and C approaches had similar trajectories, WT reached lower values earlier, with a delay between them approximating the generation interval (5 +/- 1.9 days), the rationale being described previously [17]. This led to a consistent 4-6 days difference in the median estimated time for *R*_*t*_ becoming <1, which can be seen in Figure 1 lower row for both regions. The validity of *R*_*t*_ estimates, especially using the WT method, appeared to decrease towards the latest dates.

## Discussion

We proposed a three-step approach to characterize an outbreak in near real-time by adjusting for incomplete data and reporting delays. We applied this approach to the early SARS-CoV-2 outbreak in two regions of Spain. Our findings showed that a country-wide lockdown control led to a substantial decline in cases shortly thereafter, around March 14-20 in Madrid and around March 16-21 in Murcia. In contrast, a naïve approach based on reported cases detected the decline in cases 10 days after the lockdown in Madrid and 8 after the lockdown in Murcia.

The *R*_*t*_ estimates obtained from nowcasted case counts were more consistent with true transmission (as documented using complete data that became available after the study period). For example, heterogeneity by day-of-the-week was more prominent in reported case counts than in our reconstructions for Madrid, and the expected differences between Cori’s and WT’s due to the generation interval were observed for the nowcast but not when using reported case counts day in Murcia [17].

Our approach has several limitations. First, its validity relies on the assumption that the date of symptoms onset is missing at random and that the available historical data were sufficient to parameterize the unknown reporting delay. However, our sensitivity analyses indicate that the overall trajectory of the epidemic curve was relatively robust to small departures from these assumptions. Second, our approach underperforms well when little information is available for training the nowcasting algorithm, including good estimates of the reporting delay distribution. Our estimates were sensitive to the choice of imputation and nowcasting procedures when the date of symptoms onset was unknown for a high percentage of confirmed cases. Third, our approach requires that underdetection/underreporting of cases does not change significantly over time; otherwise would adversely affect the estimates [23]. Finally, our estimates could be improved by reconstructing the epidemic curve by the date of infection rather than that of symptoms onset, though this would require more complex methods given that the temporal delay from infection to symptom onset is much harder to characterize [17,24–27].

Development of ready-to-use tools for epidemic dynamics modelling help surveillance services to appropriately present data for efficient epidemic control, but understanding the limitations of the procedure and the impact of prespecified assumptions is critical for interpretation. We believe that the contribution of our approach is twofold. First, we provide a systematic analysis on the assumptions and implementation procedures frequently used to characterize emerging outbreaks. Second, we propose a disease surveillance framework that acknowledges and adjusts for biases arising from real-world observational challenges, and is capable of providing objective, quantifiable, and systematic information, to aid the decision-making process in real-time outbreak mitigation efforts.

## Data Availability

The data that support the findings of the study was obtained from the Spanish System for Surveillance at the National Center of Epidemiology (RENAVE) through the Web platform SiViEs (System for Surveillance in Spain). Data is publicly available as aggregated daily numbers at https://cnecovid.isciii.es/covid19/.

## Acknowledgments

We thank Laura White and Rene Niehus for their technical assistance.

